# Development of highly specific singleplex and multiplex real-time reverse transcription PCR assays for the identification of SARS-CoV-2 Omicron BA.1, BA.2 and Delta variants

**DOI:** 10.1101/2022.04.07.22273168

**Authors:** Jeremy A. Garson, Samuel Badru, Anjna Badhan, Simon Dustan, Richard S. Tedder

## Abstract

The Omicron variant of SARS-CoV-2 (B.1.1.529), first identified during November 2021, is rapidly spreading throughout the world, replacing the previously dominant Delta variant. Omicron has a high number of mutations in the spike gene, some of which are associated with greatly increased transmissibility and immune evasion. The BA.1 sublineage has been most prevalent but there is recent evidence that the BA.2 sublineage is increasing in proportion in many countries. Genome sequencing is the gold standard for Omicron identification but is relatively slow, resource intensive, of limited capacity and often unavailable. We therefore developed a simple, rapid reverse transcription PCR (RT-PCR) method for sensitive and specific detection of the Omicron variant, including both the BA.1 and BA.2 sublineages. The assay targets a total of 5 nucleotide mutations in the receptor binding domain of the spike gene that give rise to 4 amino acid substitutions at G339D, S371L, S373P and S375F. The forward primer was designed as a double-mismatch allele specific primer (DMAS) with an additional artificial mismatch located four nucleotides from the 3’ end to enhance binding specificity. Assay specificity was confirmed by testing a wide range of previously-sequenced culture-derived viral isolates and clinical samples including the Alpha, Beta and Delta variants and ‘wild type’ SARS-CoV-2. Respiratory syncytial virus and influenza A were also tested. The assay can be run in singleplex format, or alternatively as a multiplex RT-PCR to enable Omicron and Delta variants to be detected and distinguished within the same reaction by means of probes labelled with different fluorescent dyes. Sublineages BA.1 and BA.2 can be differentiated if required. The methods presented here can readily be established in any PCR laboratory and should provide valuable support for epidemiologic surveillance of Omicron infections, particularly in those regions that lack extensive sequencing facilities.

## INTRODUCTION

Genomes of the causative agent of the COVID-19 global pandemic, severe acute respiratory syndrome coronavirus 2 (SARS-CoV-2), have been intensively analysed since the virus was first identified in January 2020. Since that time almost nine million genome sequences have been submitted to the GISAID open access database [GISAID, 2022]. Analysis of these sequences has tracked the evolution of SARS-CoV-2 as numerous variants have emerged throughout the world. Five of these have been designated as Variants of Concern (VoC) by the World Health Organization based on characteristics such as increased virulence, increased transmissibility and/or decreased effectiveness of vaccines, therapeutics or diagnostics [WHO, 2022]. Four of the VoC, Alpha (B.1.1.7), Beta (B.1.351), Gamma (P1) and Delta (B.1.617.2), were first identified during 2020 in the UK, South Africa, Brazil and India respectively [Davies et al., 2021; Tegally et al., 2021; Faria et al., 2021; Mlcochova et al., 2021]. Delta was dominant globally until November 2021 when the Omicron variant (B.1.1.529) emerged in multiple countries within a few weeks [Karim and Karim, 2021]. Since then, Omicron has rapidly outcompeted pre-existing variants and is replacing Delta in many parts of the world.

Omicron is characterised by an unusual constellation of mutations, with approximately 30 in the spike protein, mostly located in the N-terminal domain and receptor-binding domain. These mutations are thought to be responsible for its extremely high transmissibility and its ability to partially evade vaccine-induced immunity and immunity arising from previous SARS-CoV-2 infections, and to resist therapeutic monoclonal antibodies [Planas et al., 2022]. Fortunately, the available evidence suggests that Omicron may be associated with less severe clinical disease than other VoC and a reduced risk of a hospitalization [Ferguson et al., 2021]. Nevertheless, despite lower severity, significant morbidity and deaths are occurring, particularly in vulnerable populations, and are likely to continue with substantial pressure on health services, given the high levels of community transmission [WHO, 2022b].

Monitoring the spread of Omicron is thus important to guide public health policies and interventions. Although genome sequencing is regarded as the gold standard for Omicron identification it is relatively expensive, complex, slow (days/weeks turnaround time), resource intensive, and unavailable in many regions [Brito et al., 2021]. Alternative techniques offering rapid, high-throughput, inexpensive genotyping for efficient surveillance of Omicron are therefore desirable. To address this need, we developed a highly specific real-time reverse transcription PCR (RT-PCR), using a similar design strategy to that employed in our recently published Delta variant RT-PCR genotyping assay [Garson et al., 2022]. Specificity was confirmed by testing a range of previously-sequenced culture-derived viral isolates and clinical samples.

Until January 2022, almost all Omicron infections were due to the BA.1 sublineage but reports from Denmark, India, the UK and elsewhere suggest that the BA.2 sublineage is increasing in proportion and may shortly overtake BA.1 [Chen and Wei, 2022; WHO 2022c]. BA.2 has a number of unique mutations in the spike gene, several of which involve the target region of the Omicron specific RT-PCR described in this study. In order to accommodate these BA.2 mutations a modified reverse primer was designed. The assay format is flexible and can be used to detect the BA.1 and BA.2 sublineages separately or in combination. In addition, we demonstrate that this Omicron-specific assay can be multiplexed effectively with the Delta-specific assay previously reported [Garson et al., 2022].

## MATERIALS AND METHODS

### Cell cultured viral isolates

SARS-CoV-2 RNA extracted from Vero cell supernatants was kindly provided by Professor Wendy Barclay, Department of Infectious Disease, Imperial College London. RNA was aliquoted and stored at -80°C. The following isolates were obtained: Omicron #O1 and #O2 are B.1.1.529 sublineage BA.1 isolates designated M21021166 and NWLP04 respectively. IC19 (hCoV-19/England/IC19/2020|EPI_ISL_475572|2020-03-17), a B.1 lineage virus with the D614G spike mutation but otherwise identical to the original “wild-type” Wuhan virus. Alpha #246 (hCoV-19/England/205080610/2020|EPI_ISL_723001), an example of the B.1.1.7 Alpha variant. Beta #65 (hCoV-19/England/205280030/2020|EPI_ISL_770441|2020-12-24) and Beta #78 are examples of the B.1.351 Beta variant. Delta #395 and Delta #02510 are examples of the B.1.617.2 Delta variant. The identities of all these viral isolates had been confirmed by full genome sequencing. Influenza A virus PR8 (A/Puerto Rico/8/1934(H1N1) RNA was also provided by Professor Barclay and human respiratory syncytial virus (RSV) strains PP3L and PP3KL were obtained from Dr John Tregoning, Department of Infectious Disease, Imperial College London.

### Clinical samples

Residual, anonymised nasopharyngeal and oropharyngeal swab samples in virus transport medium were obtained from staff and students at Imperial College London, and from household contacts of individuals with COVID-19 (Research Ethics Committee reference 20/NW/0231, IRAS ID: 282820). Extracted RNA from the samples was stored at -80°C for between 2 weeks and 5 months before being used in the present study. Initial screening of clinical samples for SARS-CoV-2 had been performed using a duplex RT-qPCR assay which targets both the E gene (Charité assay) and a human RNA transcript, RNase P (CDC assay) as an internal sample sufficiency control [Rowan et al., 2021].

### RNA extraction

Viral RNA was extracted from clinical samples with a CyBio Felix liquid handing robot (Analytik Jena) and InnuPREP Virus DNA/RNA Kit (Analytik Jena), used according to manufacturer’s instructions. RNA was eluted in 50 μL of RNase-free water and stored at -80°C until use.

### Sequencing to establish viral genotype

Clinical samples in which SARS-CoV-2 had been detected were sequenced at the Molecular Diagnostics Unit (MDU), Department of Infectious Disease, Imperial College London. Libraries were amplified using the EasySeq(tm) RC-PCR SARS CoV-2 (novel coronavirus) WGS kit (NimaGen) and run on an Illumina iSeq 100 next-generation sequencing system.

### Primer and probe design

The Omicron variant has an unusually high number of mutations, approximately 30, in the gene encoding the spike protein. Although many of these individual mutations are shared by other variants, several of them such as S371L, appear to be unique. Using alignments, performed by MEGA version 7.0.21, of SARS-CoV-2 spike gene sequences downloaded from the GISAID database (https://www.gisaid.org/), we searched for combinations of spike mutations which would permit the Omicron variant to be differentiated from all other variants of concern and from ‘wild type’ SARS-CoV-2. We noted a cluster of 5 nucleotide mutations in the receptor binding domain (RBD) that encoded 4 amino acid substitutions, at G339D, S371L, S373P and S375F. All of these mutations were well conserved in a set of 58 downloaded Omicron sequences originating from diverse countries including the Netherlands, England, Scotland and South Africa. By incorporating G339D in the forward primer and S371L, S373P and S375F in the reverse primer we were able to differentiate Omicron from the other VoC *in silico*. The forward primer was designed with an additional artificial internal mismatch, four bases from the 3’ end. This design, referred to as a double-mismatch allele-specific primer (DMAS) was first described by Lefever [Lefever et al., 2019] and has been shown to significantly enhance binding specificity. The reverse primer has 4 natural mismatches and therefore requires no additional artificial mismatch to achieve high specificity. The primer pair generates an amplicon of 146 bp and is detected by a fluorescently-labelled hydrolysis probe.

Infections due to the BA.2 sublineage of Omicron were recognised as numerically significant in certain countries after this primer pair and probe were designed. Sequence analysis of 20 BA.2 sequences downloaded from the GISAID database revealed that although the forward primer and the probe would detect both BA.1 and BA.2 sublineages, the reverse primer would probably not amplify BA.2 efficiently due to two nucleotide differences between BA.1 and BA.2 at this location. We therefore designed a modified reverse primer to match the BA.2 sequence. This can be used in combination with the BA.1 reverse primer to detect both sublineages, or in a separate reaction if distinguishing between BA.1 and BA.2 is required.

Primers (Table 1) were checked by *in silico* PCR (https://genome.ucsc.edu/cgi-bin/hgPcr) to rule out cross reactivity with the human genome, and by NCBI BLASTn (https://blast.ncbi.nlm.nih.gov/Blast.cg) to exclude reactivity with other respiratory viruses including human coronaviruses 229E, OC43 and NL63. Melting temperature (Tm) estimations and checks for primer dimers or significant secondary structure were also performed (IDT OligoAnalyzer, https://eu.idtdna.com/calc/analyzer).

**Table 1.**
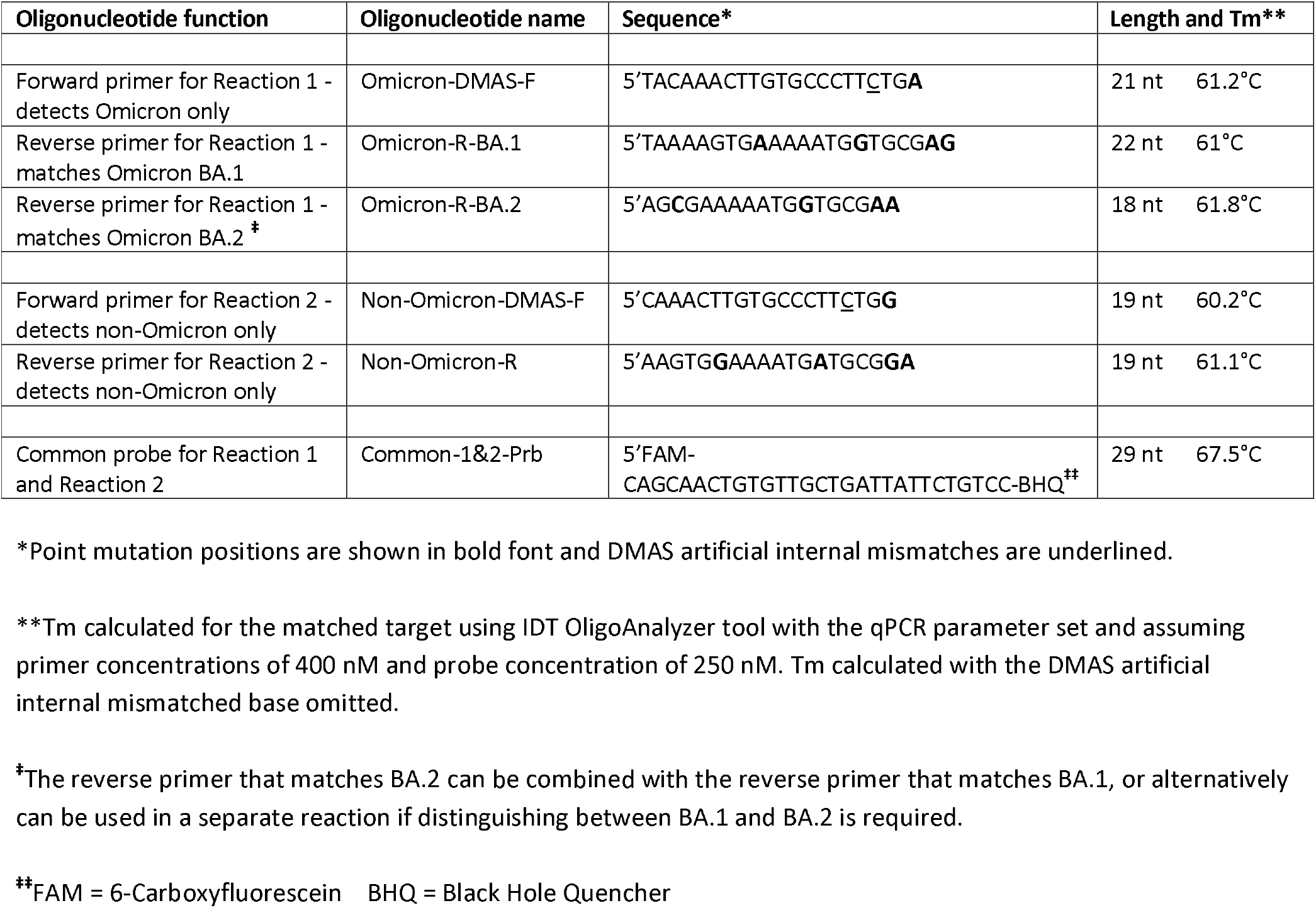
Details of primer and probe sequences used for the Omicron RT-PCR assay.

### RT-PCR singleplex assay to determine whether a SARS-CoV-2 sample is the Omicron variant

The method requires two parallel RT-PCR reactions to be carried out in separate wells. Reaction 1 is designed to specifically detect Omicron only and Reaction 2 is designed to detect all SARS-CoV-2 variants other than Omicron. Primer sequences and the sequence of the fluorescently labelled hydrolysis probe are detailed in Table 1. Primers and probes were synthesized by Integrated DNA Technologies (IDT, Belgium). Five μL RNA template were used in a 20 μL reaction containing 5 μL of 4x TaqMan Fast virus 1-step mastermix (Applied Biosystems), primers at 400 nM and probe at 250 nM. Thermal cycling was performed in a Bio-Rad CFX real-time PCR system with reverse transcription at 54°C for 10 min, followed by 94°C for 3 min, then 40 cycles of 94°C for 15 sec and 58°C for 30 sec. Data were processed using Bio-Rad CFX maestro 2.0 software with baseline subtracted curve fit, fluorescence drift correction, automatically calculated baseline cycles and manual threshold settings. Each run included Omicron #O1 RNA and Delta #395 RNA, both diluted a million-fold, as positive controls. No-template nuclease-free water negative controls were also included in each run. As the assay is designed primarily for genotyping rather than for quantification it does not require a calibration curve. If a test sample generates an amplification curve in Reaction 1 but not in Reaction 2 it is interpreted as ‘Omicron detected’. Conversely, if a test sample generates an amplification curve in Reaction 2 but not in Reaction 1, it is interpreted as ‘Non-Omicron SARS-CoV-2 detected’. If neither reaction produces an amplification curve, the interpretation is ‘SARS-CoV-2 not detected’.

### Multiplex Omicron and Delta RT-PCR assay

In many parts of the world the only two VoC likely to be in circulation at the present time are Omicron and to a greater or lesser extent, Delta. We therefore designed a multiplex (duplex) RT-PCR assay able to detect both Omicron and Delta within the same reaction well. This employed a combination of the primer/probe set described above for Reaction 1 (Omicron-specific) and the Delta-specific primer/probe set previously published [Garson et al, 2022]. To be able to differentiate between the signals generated, the fluorescent probe for the Delta variant was labelled with a different fluorescent dye (HEX) (Eurogentec, Belgium) from that used to label the Omicron probe (FAM). Sequence details of the Delta primer/probe set are as follows: forward primer, designated Delta_DMAS_F 5’GGTTGGTGGTAATTATAATTCCCG; reverse primer, designated Delta_DMAS_R 5’CCTTCAACACCATTACAACGTT; probe, designated Delta_Prb 5’HEX-TCTCTCAAAAGGTTTGAGATTAGACTTCC-BHQ. The mastermix, primer and probe concentrations and the thermal cycling parameters are as described above for the Omicron RT-PCR singleplex assay.

## RESULTS

### Cell cultured viral isolates

Ten-fold dilution series of viral RNA extracted from cell culture supernatants were prepared, starting at a dilution of 1:1,000. The dilution series were tested in Reaction 1 and Reaction 2 as described above under Materials and Methods. Figure 1 illustrates typical amplification curves generated by Reaction 1, designed to be Omicron-specific, with a dilution series of Omicron variant RNA (#O1 M21021166). The Omicron RNA remains detectable at a dilution of 1:1,000,000 whereas no amplification occurs with non-Omicron variants such as Delta, even at the lowest 1:1,000 dilution tested. Almost identical results were found by testing dilution series of the second Omicron isolate (#O2 NWLP04) with Reaction 1. Conversely, Reaction 2 (designed to detect non-Omicron SARS-CoV-2 only) generated no signal with either Omicron isolate at any dilution. As expected, Reaction 2 detected all non-Omicron variants tested, IC19, Alpha #246, Beta #65 and Beta #78. These results are summarised in Table 2.

**Table 2.** RT-PCR assay results on cell cultured SARS-CoV-2 isolates.

| Viral isolate | Sequencing result | Reaction 1 for Omicron. Highest dilution detectable** | Reaction 2 for non-Omicron. Highest dilution detectable** | Genotype by RT-PCR genotyping assay |
| --- | --- | --- | --- | --- |
| #O1 M21021166 | Omicron BA.1 | 1:1,000,000 | Not detected at any dilution | Omicron variant |
| #O2 NWLP04 | Omicron BA.1 | 1:10,000,000 | Not detected at any dilution | Omicron variant |
| Alpha #246 | Alpha variant | Not detected at any dilution | 1:100,000 | Not Omicron variant |
| Beta #65 | Beta variant | Not detected at any dilution | 1:1,000,000 | Not Omicron variant |
| Beta #78 | Beta variant | Not detected at any dilution | 1:1,000,000 | Not Omicron variant |
| Delta #395 | Delta variant | Not detected at any dilution | 1:1,000,000 | Not Omicron variant |
| Delta #02510 | Delta variant | Not detected at any dilution | 1:10,000,000 | Not Omicron variant |
| IC19 | 'wild type'* | Not detected at any dilution | 1:1,000,000 | Not Omicron variant |

\*IC19 is a B.1 lineage SARS-CoV-2 similar to the original 'wild-type' Wuhan virus but with the D614G spike mutation.
\*\*Ten-fold dilution series started at 1:1,000

**Figure 1.**
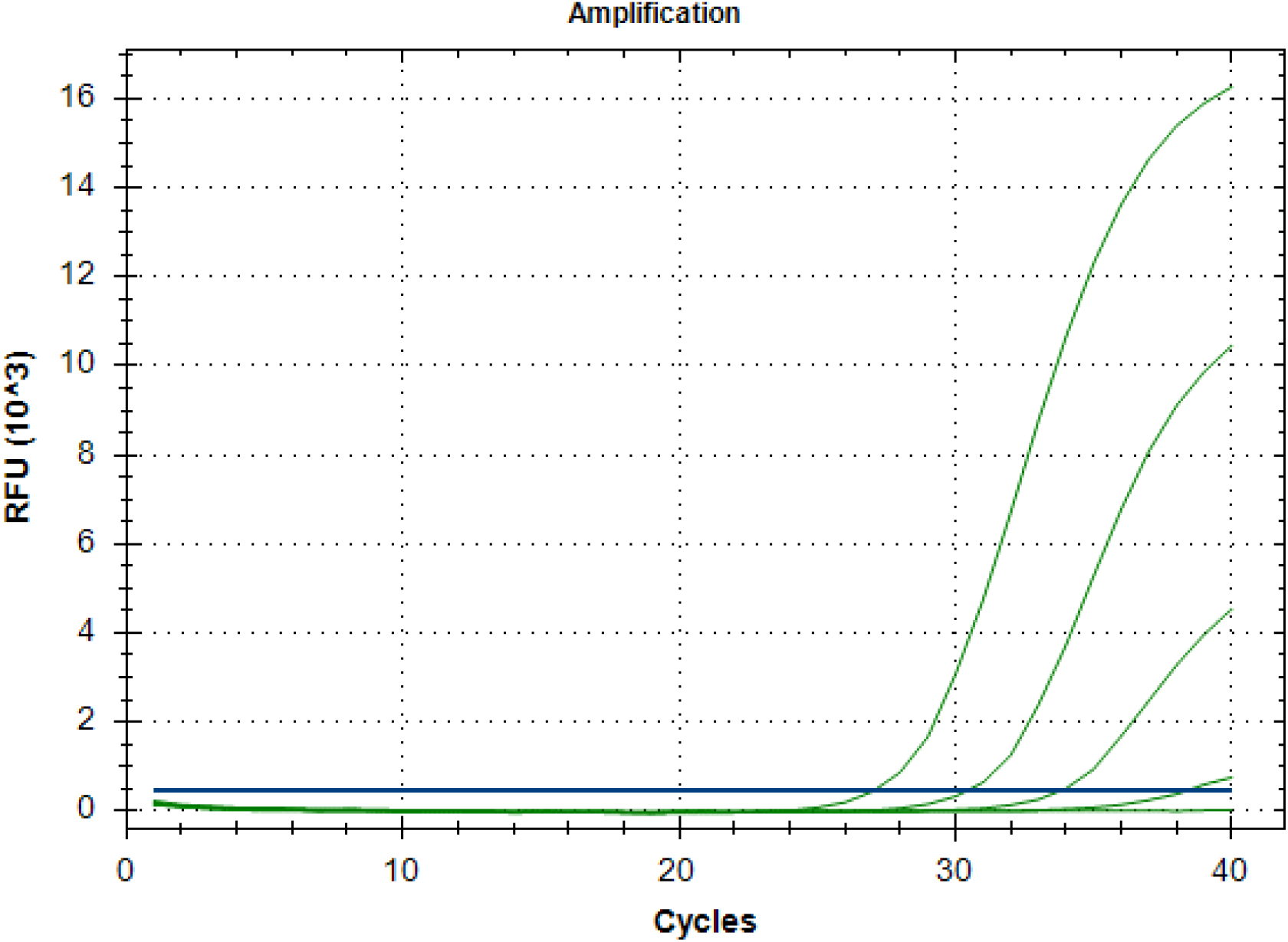
Typical amplification curves generated by Reaction 1 with dilution series of Omicron RNA. A ten-fold dilution series of Omicron variant RNA (#O1 M21021166) was amplified by Reaction 1. From the left the four green amplification curves represent the signals from dilutions at 1:1,000, 1:10,000, 1:100,000 and 1:1,000,000 respectively. The threshold level is shown as the horizontal dark blue line which is crossed by all four amplification curves. The horizontal green line below the threshold represents non amplification of the Delta variant (Delta #02510) at 1:1,000 dilution.

### Clinical samples

Results from testing clinical samples that had been genotyped by sequencing, paralleled the results generated by testing sequence-confirmed cultured viral isolates. Thus, clinical samples that had been shown to be Omicron by genome sequencing (n = 16) or presumed to be Omicron by virtue of the date that they were taken (n = 5), were detected by Reaction 1 but not by Reaction 2 and conversely, clinical samples that had been shown to be non-Omicron variants by sequencing were detected by Reaction 2 but not by Reaction 1 (Table 3). In the 5 cases where sequencing had either failed or was not performed, the samples were expected to be Omicron because they were taken during early March 2022 when approximately 99.7% of UK SARS-CoV-2 samples were due to the Omicron variant [UKHSA, 2022]. All tested clinical samples in which SARS-CoV-2 had been detected by the E gene screening assay [Rowan et al., 2021] were successfully genotyped by the singleplex RT-PCR assay. There was no evidence of false-positive results being generated by either Reaction 1 or Reaction 2; no-template controls were consistently negative as were 12 clinical samples that had tested negative for SARS-CoV-2 in the E gene PCR screening assay.

**Table 3.** RT-PCR singleplex results show concordance with genotype determined by sequencing of clinical samples.

| Sample No. | Genotype by sequencing | Reaction 1 Ct (Omicron) | Reaction 2 Ct (non-Omicron) | Genotype by RT-PCR | Comments |
| --- | --- | --- | --- | --- | --- |
| SB4 | Delta | Not detected‡ | 22.45 | Not Omicron | Concordant result |
| SB5 | Delta | Not detected‡ | 39.04 | Not Omicron | Concordant result |
| SB6 | Delta | Not detected‡ | 27.86 | Not Omicron | Concordant result |
| SB7 | Delta | Not detected‡ | 26.30 | Not Omicron | Concordant result |
| SB8 | Delta | Not detected‡ | 29.43 | Not Omicron | Concordant result |
| SB36 | Omicron BA.1 | 28.73* | Not detected | Omicron | Concordant result |
| SB37 | Omicron BA.1 | 31.03* | Not detected | Omicron | Concordant result |
| SB38 | Omicron BA.1 | 25.06* | Not detected | Omicron | Concordant result |
| SB39 | Omicron BA.1 | 30.93* | Not detected | Omicron | Concordant result |
| SB40 | Omicron BA.1 | 31.10* | Not detected | Omicron | Concordant result |
| SB41 | Omicron BA.1 | 32.43* | Not detected | Omicron | Concordant result |
| SB42 | Omicron BA.1 | 35.15* | Not detected | Omicron | Concordant result |
| SB43 | Omicron BA.1 | 32.74* | Not detected | Omicron | Concordant result |
| SB44 | Omicron BA.1 | 35.89* | Not detected | Omicron | Concordant result |
| SB45 | Omicron BA.1 | 25.38* | Not detected | Omicron | Concordant result |
| SB46 | Omicron BA.1 | 28.08* | Not detected | Omicron | Concordant result |
| SB47 | Omicron BA.2 | 28.24† | Not detected | Omicron | Concordant result |
| SB49 | Omicron BA.2 | 24.98† | Not detected | Omicron | Concordant result |
| SB53 | Omicron BA.2 | 23.31† | Not detected | Omicron | Concordant result |
| SB55 | Omicron BA.2 | 31.81† | Not detected | Omicron | Concordant result |
| SB56 | Omicron BA.2 | 34.11† | Not detected | Omicron | Concordant result |
| SB48 | Unknown<sup>§</sup> | 35.47† | Not detected | Omicron | Expected to be Omicron†† |
| SB50 | Sequencing failed | 31.62† | Not detected | Omicron | Expected to be Omicron†† |
| SB52 | Unknown<sup>§</sup> | 33.92† | Not detected | Omicron | Expected to be Omicron†† |
| SB54 | Unknown<sup>§</sup> | 37.72† | Not detected | Omicron | Expected to be Omicron†† |
| SB57 | Unknown<sup>§</sup> | 36.93† | Not detected | Omicron | Expected to be Omicron†† |

\*Reaction 1 was performed with reverse primer Omicron-R-BA.1
†Reaction 1 was performed with reverse primer Omicron-R-BA.2
‡Reaction 1 was performed with a 1:1 mixture of reverse primers Omicron-R-BA.1 and Omicron-R-BA.2
<sup>§</sup>Unknown because this sample was not sent for sequencing
††Expected to be Omicron because ~ 99.7% of UK SARS-CoV-2 samples were Omicron during early March 2022 when the samples were taken [UKHSA, 2022]

Furthermore, it was possible to determine whether an Omicron sample belonged to sublineage BA.1 or BA.2 by comparing the Ct (cycle threshold) values generated by running the BA.1 reverse primer and the BA.2 reverse primer in separate reactions. Sequence-confirmed BA.1 samples always generated earlier/lower Ct values with the BA.1 reverse primer reaction and conversely, BA.2 samples always generated earlier/lower Ct values with the BA.2 reverse primer reaction. In all cases the designation of sublineage by RT-PCR agreed with the sublineage determined by genome sequencing (Table 4).

**Table 4.** Omicron sublineage determined by RT-PCR agrees with sublineage determined by genome sequencing.

| **Sample No.** | **Sublineage by sequencing** | **Reaction 1 Ct with primer Omicron-R-BA.1** | **Reaction 1 Ct with primer Omicron-R-BA.2** | **Sublineage by RT-PCR** | **Comments** |
| --- | --- | --- | --- | --- | --- |
| SB12 | BA.2 | 25.15 | 24.45 | BA.2 | Sublineages concur |
| SB13 | BA.2 | 33.71 | 32.91 | BA.2 | Sublineages concur |
| SB14 | BA.1 | 26.85 | 32.08 | BA.1 | Sublineages concur |
| SB15 | BA.1 | 25.77 | 31.71 | BA.1 | Sublineages concur |
| SB16 | BA.1 | 27.77 | 33.09 | BA.1 | Sublineages concur |
| SB17 | BA.2 | 34.94 | 33.22 | BA.2 | Sublineages concur |
| SB18 | BA.2 | 25.10 | 23.68 | BA.2 | Sublineages concur |
| SB19 | BA.2 | 29.10 | 28.47 | BA.2 | Sublineages concur |
| SB20 | BA.1 | 28.75 | 34.71 | BA.1 | Sublineages concur |
| SB21 | BA.1 | 29.15 | 35.12 | BA.1 | Sublineages concur |

### Multiplex RT-PCR assay

The multiplex RT-PCR assay, designed to detect Omicron and Delta variants within the same well, performed as expected. No reduction in sensitivity in comparison with the component singleplex assays was observed in testing dilution series of cell cultured viral isolates O1 (M21021166), O2 (NWLP04), Delta #395 and Delta #02510. As predicted, the other SARS-CoV-2 viral isolates IC19, Alpha #246, Beta #65 and Beta #78 generated no signal in the multiplex assay.

The results of multiplex RT-PCR testing of clinical samples are presented in Table 5. Thirty four samples were tested including 10 that were sequence-confirmed Delta and one that was expected to be Delta by virtue of the date that the sample was taken (early October 2021). In all but one of these 11 cases the multiplex assay indicated that the Delta variant was present in the sample. In the remaining case both the Omicron and Delta reactions were negative, due to extremely low titre RNA (Ct value 39.04 in the corresponding singleplex assay), causing failure to amplify as a result of stochastic effects. The clinical samples tested also included 23 sequence-confirmed Omicron variants, both BA.1 and BA.2 sublineages. The multiplex assay confirmed the presence of Omicron in all but two of these samples, both having extremely low viral RNA titres with detection therefore subject to stochastic variability.

**Table 5.**
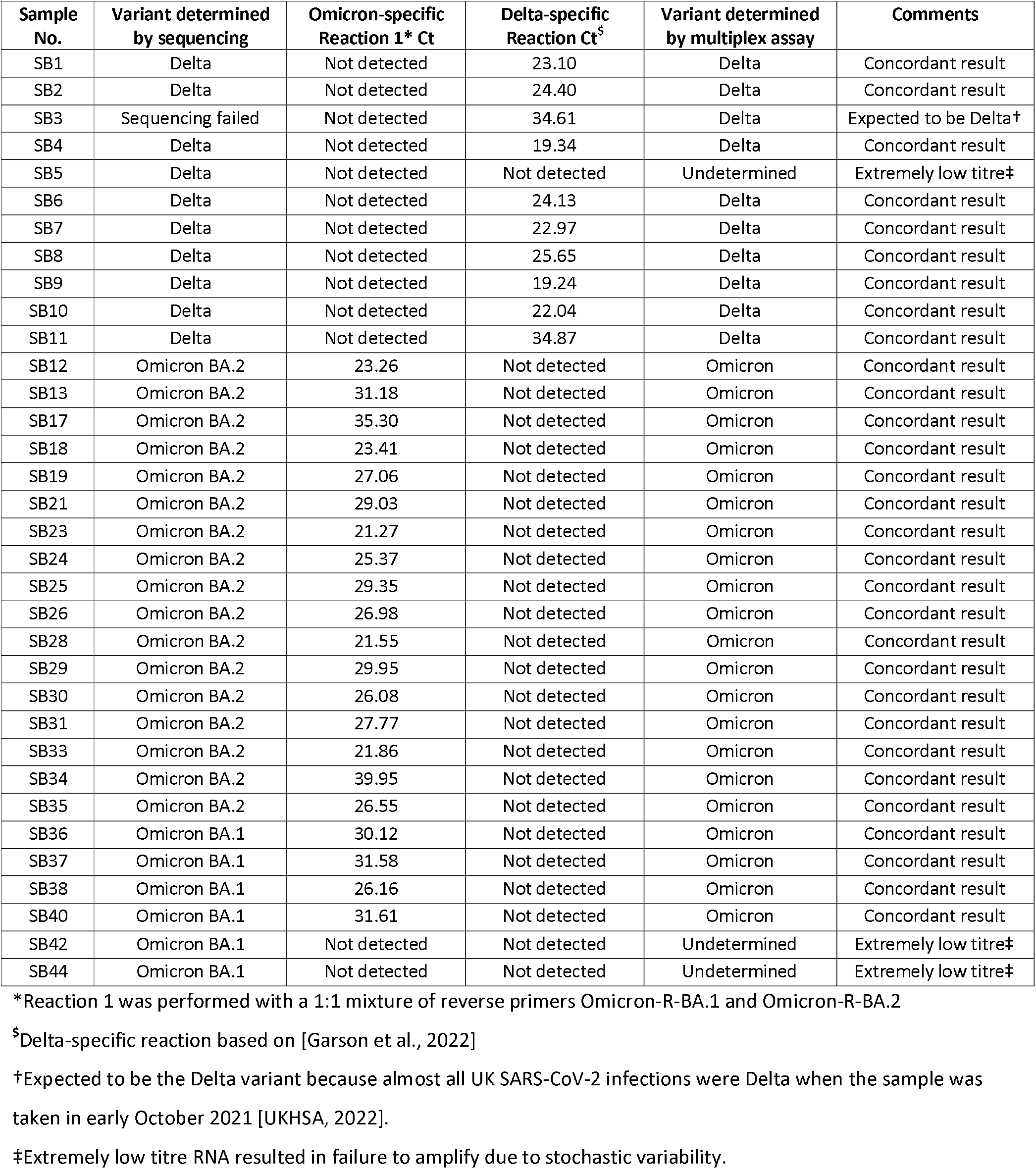
Omicron/Delta multiplex RT-PCR assay testing of previously sequenced clinical samples.

| Sample No. | Variant determined by sequencing | Omicron-specific Reaction 1* Ct | Delta-specific Reaction Ct<sup>§</sup> | Variant determined by multiplex assay | Comments |
| --- | --- | --- | --- | --- | --- |
| SB1 | Delta | Not detected | 23.10 | Delta | Concordant result |
| SB2 | Delta | Not detected | 24.40 | Delta | Concordant result |
| SB3 | Sequencing failed | Not detected | 34.61 | Delta | Expected to be Delta† |
| SB4 | Delta | Not detected | 19.34 | Delta | Concordant result |
| SB5 | Delta | Not detected | Not detected | Undetermined | Extremely low titre‡ |
| SB6 | Delta | Not detected | 24.13 | Delta | Concordant result |
| SB7 | Delta | Not detected | 22.97 | Delta | Concordant result |
| SB8 | Delta | Not detected | 25.65 | Delta | Concordant result |
| SB9 | Delta | Not detected | 19.24 | Delta | Concordant result |
| SB10 | Delta | Not detected | 22.04 | Delta | Concordant result |
| SB11 | Delta | Not detected | 34.87 | Delta | Concordant result |
| SB12 | Omicron BA.2 | 23.26 | Not detected | Omicron | Concordant result |
| SB13 | Omicron BA.2 | 31.18 | Not detected | Omicron | Concordant result |
| SB17 | Omicron BA.2 | 35.30 | Not detected | Omicron | Concordant result |
| SB18 | Omicron BA.2 | 23.41 | Not detected | Omicron | Concordant result |
| SB19 | Omicron BA.2 | 27.06 | Not detected | Omicron | Concordant result |
| SB21 | Omicron BA.2 | 29.03 | Not detected | Omicron | Concordant result |
| SB23 | Omicron BA.2 | 21.27 | Not detected | Omicron | Concordant result |
| SB24 | Omicron BA.2 | 25.37 | Not detected | Omicron | Concordant result |
| SB25 | Omicron BA.2 | 29.35 | Not detected | Omicron | Concordant result |
| SB26 | Omicron BA.2 | 26.98 | Not detected | Omicron | Concordant result |
| SB28 | Omicron BA.2 | 21.55 | Not detected | Omicron | Concordant result |
| SB29 | Omicron BA.2 | 29.95 | Not detected | Omicron | Concordant result |
| SB30 | Omicron BA.2 | 26.08 | Not detected | Omicron | Concordant result |
| SB31 | Omicron BA.2 | 27.77 | Not detected | Omicron | Concordant result |
| SB33 | Omicron BA.2 | 21.86 | Not detected | Omicron | Concordant result |
| SB34 | Omicron BA.2 | 39.95 | Not detected | Omicron | Concordant result |
| SB35 | Omicron BA.2 | 26.55 | Not detected | Omicron | Concordant result |
| SB36 | Omicron BA.1 | 30.12 | Not detected | Omicron | Concordant result |
| SB37 | Omicron BA.1 | 31.58 | Not detected | Omicron | Concordant result |
| SB38 | Omicron BA.1 | 26.16 | Not detected | Omicron | Concordant result |
| SB40 | Omicron BA.1 | 31.61 | Not detected | Omicron | Concordant result |
| SB42 | Omicron BA.1 | Not detected | Not detected | Undetermined | Extremely low titre‡ |
| SB44 | Omicron BA.1 | Not detected | Not detected | Undetermined | Extremely low titre‡ |

\*Reaction 1 was performed with a 1:1 mixture of reverse primers Omicron-R-BA.1 and Omicron-R-BA.2
<sup>§</sup>Delta-specific reaction based on [Garson et al., 2022]
†Expected to be the Delta variant because almost all UK SARS-CoV-2 infections were Delta when the sample was taken in early October 2021 [UKHSA, 2022].
‡Extremely low titre RNA resulted in failure to amplify due to stochastic variability.

### Specificity of the RT-PCR assays

The high specificity of the assays permitted them to differentiate reliably between the Omicron variant and other non-Omicron variants of SARS-CoV-2. The BA.1 and BA.2 sublineages could also be distinguished from each other as described above. Respiratory syncytial virus and influenza A were selected as examples of other common respiratory viruses that it would be essential for these RT-PCR assays not to detect. We were able to confirm that the RT-PCR assays did not generate any non-specific signal with either Influenza A virus PR8 or two RSV strains, PP3L and PP3KL.

## DISCUSSION

The RT-PCR method described here, which uses a combination of a double-mismatch allele-specific forward primers (DMAS) and conventional allele-specific reverse primers, has been shown in this study to be capable of reliably detecting the Omicron variant and differentiating it from other non-Omicron SARS-CoV-2 variants. It is also able to differentiate between the two major Omicron sublineages BA.1 and BA.2. By employing this method, which should be easy to establish in any laboratory conducting PCR assays, the Omicron variant can be identified with confidence without having to resort to relatively complex, expensive and time-consuming genome sequencing. Even in those well-resourced countries where genome sequencing is available, the magnitude of the recent surge in cases of Omicron threatens to overwhelm sequencing capacity and result in delayed reporting of variants. The availability of rapid and relatively inexpensive genotyping techniques such as that described in this study should greatly facilitate epidemiological surveillance throughout the world.

Failure of the S-gene to amplify in certain commercial assays, such as the Applied Biosystems TaqPath Covid-19 PCR test, due to the 69-70del spike mutation, was used initially as a proxy marker for Omicron by some laboratories [Karim and Karim, 2021]. However, although this S-gene target failure (SGTF) method was relatively simple, it has proven increasingly unreliable, particularly since the emergence of the BA.2 sublineage which lacks the 69-70del mutation [UKHSA, 2022]. The inability of BA.2 to be identified by the SGTF method has led to it being described as ‘Stealth Omicron’ [Christensen et al., 2022]. The RT-PCR genotyping method we describe here does not have this disadvantage and is capable of detecting both BA.1 and BA.2 sublineages reliably.

Depending on the relative proportions of Omicron and other variants in a given location at any particular time, it may be more appropriate to run the assays in singleplex format, either to differentiate Omicron from other variants and/or to discriminate between the BA.1 and BA.2 sublineages. Indeed, the WHO Technical Advisory Group on SARS-CoV-2 Virus Evolution advised that, “BA.2 should continue to be monitored as a distinct sublineage of Omicron by public health authorities” [WHO, 2022c].

The testing strategy can be flexible so that in areas where the Delta variant is still present it may be more efficient to use the multiplex format to provide data on the relative prevalence of Delta and Omicron in the population. On the other hand, in countries such as the UK where the Omicron variant already represents the vast majority of cases, it may be more helpful to use the singleplex assays to identify the small minority of samples which are not Omicron and to select these for sequencing in order to maximise the chance of early discovery of novel, emergent non-Omicron variants.

## Data Availability

All data produced in the present work are contained in the manuscript

## ACKNOWLEDGMENTS

We are grateful to the NIHR BRC at Imperial College Healthcare Trust for its support of this study. We thank Professor Wendy Barclay and Dr John Tregoning for kindly providing viral isolates, and Professors Graham Taylor and Ajit Lalvani for their support. We are also grateful to Professor Myra McClure for her invaluable support and encouragement.

## AUTHOR CONTRIBUTIONS

Conceptualisation, JAG, RST; Investigation, SB, AB, SD; Methodology, JAG, SB; Project administration, JAG, RST; Supervision, JAG; Writing – original draft, JAG; Writing – review & editing, JAG, SB, AB, RST; Final approval of submitted version, JAG, SB, RST, AB, SD.

